# Prescribing of GLP-1 Receptor Agonists for Adolescents with Obesity and Associated Disparities

**DOI:** 10.1101/2025.10.06.25337411

**Authors:** Chungsoo Kim, Mona Sharifi, Joseph S. Ross, Yong Chen, Hua Xu, Harlan M. Krumholz, Yuan Lu

**Affiliations:** Center for Outcomes Research and Evaluation, Yale New Haven Hospital, New Haven, Connecticut; Section of Cardiovascular Medicine, Department of Internal Medicine, Yale School of Medicine, New Haven, Connecticut; Section of General Pediatrics, Department of Pediatrics, Yale School of Medicine, New Haven, Connecticut; Department of Biomedical Informatics and Data Science, Yale School of Medicine, New Haven, Connecticut; Section of General Internal Medicine, Department of Internal Medicine, Yale School of Medicine, New Haven, Connecticut; Department of Health Policy and Management, Yale School of Public Health, New Haven, Connecticut; The Center for Health AI and Synthesis of Evidence (CHASE), Perelman School of Medicine, The University of Pennsylvania, Philadelphia; Department of Biostatistics, Epidemiology and Informatics, Perelman School of Medicine, The University of Pennsylvania, Philadelphia

## Abstract

This retrospective cohort study examined national prescribing patterns of glucagon like peptide 1 receptor agonists (GLP1RAs) for U.S. adolescents with obesity using electronic health record data from over 2 million patients aged 12 to 17 years between 2021 and 2025. Among eligible adolescents, only 0.9 received a GLP1RA prescription, though use increased after semaglutide approval in December 2022, with semaglutide rapidly surpassing liraglutide and off-label tirzepatide use rising by 2025. Prescribing was strongly associated with clinical and sociodemographic factors: adolescents with severe obesity were more likely to be prescribed, while males, Hispanic/Latino and non Hispanic Black youth, non English/Spanish speakers, those living in rural or socioeconomically disadvantaged areas, and those with Medicaid or self pay coverage were significantly less likely to receive prescriptions. These findings highlight growing uptake of GLP1RAs but reveal substantial disparities in access, suggesting insurance barriers and structural inequities may limit availability for groups disproportionately affected by obesity.

## Introduction

Glucagon-like peptide-1 receptor agonists (GLP-1RAs) achieve meaningful weight loss in adolescents with obesity.^1^ Although semaglutide was approved for adolescents in December 2022, it remains unclear which adolescents are prescribed these therapies and whether access differs by demographic and socioeconomic factors. Prior studies have described eligibility and dispensing,^2,3^ but not prescribing at the point of care. Using a large, national, electronic health record dataset, we examined GLP-1RA prescribing patterns among U.S. adolescents with obesity, including by demographic characteristics and socioeconomic status.

## Methods

We conducted a retrospective cohort study using the Epic Cosmos dataset from January 1, 2021, through July 31, 2025.^4^ Eligible patients were adolescents aged 12 to 17 years with obesity, defined as body mass index (BMI) at or above the 95th percentile for sex- and age-based growth charts by the U.S. Centers for Disease Control and Prevention. We excluded patients with a diagnosis of type 1 or type 2 diabetes. We determined whether any received a prescription for a GLP-1RA: liraglutide, semaglutide, and tirzepatide.^1^ Although not approved for adolescent obesity, we included tirzepatide to capture off-label prescribing. We excluded prescriptions for liraglutide combined with insulin.

Patient characteristics included demographics, obesity class, preferred language, insurance type, neighborhood social vulnerability, and urbanicity. We calculated the monthly incident and prevalent prescription rate, with incident prescription rates stratified by ingredient, obesity class, and insurance. Adjusted odds ratios (aORs) and 95% CIs for GLP-1RA prescribing were estimated using multivariable logistic regression models adjusting for aforementioned characteristics. This study followed the STROBE guideline and was exempt from ethics review and informed consent because it used deidentified data.

## Results

Among 2,090,467 adolescents with obesity, 19,097 (0.9%) received at least one prescription for a GLP-1 RA. Their mean (SD) age those prescribed was 15.0 (1.7) years, and 87.4% (n=16,690) had severe obesity. From the approval of semaglutide in December 2022 through July 2025, the prevalent prescription rate of GLP-1RAs increased from 0.12% to 1.38%, the incident prescription rate increased from 0.15% to 0.77%. Semaglutide rapidly surpassing liraglutide in uptake and off-label prescribing of tirzepatide has exceeded liraglutide prescribing since early 2025.

After adjustment through a multivariable model, prescribing varied across demographics and socioeconomic groups (Table). Compared with female, males were less likely to receive a prescription (aOR 0.53 [95% CI, 0.51–0.54]). Compared with non-Hispanic White adolescents, Hispanic/Latino and non-Hispanic Black adolescents had lower odds of prescriptions (aOR 0.93[0.89–0.98] and 0.89 [0.86–0.93], respectively). Adolescents in obesity class 3 were more likely to be prescribed that those in class 1 (aOR 21.19 [20.33-22.10]). Youth whose primary language was neither English nor Spanish (e.g., Chinese and Arabic) versus English also had lower odds (aOR 0.83[0.73–0.94]). Households in the most disadvantaged neighborhoods (aOR 0.61[0.58–0.64]) and those living in rural areas (aOR 0.80 [0.73–0.88]) were less likely to receive a prescription. Medicaid (aOR 0.57 [0.55–0.59]) and self-pay (0.20 [0.17–0.25]) were associated with lower odds compared with commercial insurance.

## Discussion

In this national cohort of over 2 million adolescents with obesity, prescribing of GLP-1RAs increased over time but remained below 1% of eligible adolescents. Moreover, there were disparities in prescribing, as males, Hispanic/Latino and non-Hispanic Black adolescents, those living in socioeconomically disadvantaged or rural areas, and patients insured by Medicaid were significantly less likely to receive these therapies. These disparities may reflect differences in patient or parent preferences, affordability and insurance coverage. Collectively, these factors suggest that access to GLP-1RAs is most limited among groups already disproportionately affected by obesity and adverse obesity-related outcomes.

While dispensing analyses have suggested that Medicaid beneficiaries represent a large share of GLP-1RA users in the previous study,^3^ our denominator-based results demonstrate Medicaid-covered youth were prescribed these medication at markedly lower rates. This divergence underscores the potential influence of insurance coverage restrictions and prior authorization requirements on prescribing practices. Limitations include the lack of information on prescription indication, insurance authorization outcomes, and medication adherence.

## Data Availability

All data produced in the present study are available upon reasonable request to the authors

---

**Table 1.** Demographic and Socioeconomic status of Adolescents Eligible for and Prescribed GLP-1RAs for obesity.

| Characteristics | Adolescents with obesity<br>(n = 2 090 467) | Adolescents with GLP-1RA prescription<br>(n =19 097) | Proportion (%) |
| --- | --- | --- | --- |
| Age at baseline, mean (SD) | 14.2 (1.9) | 15.0 (1.7) | - |
| Sex, n (%) |  |  |  |
| Female | 981 212 (46.9) | 11 825 (61.9) | 1.2 |
| Male | 1 109 255 (53.1) | 7 272 (38.1) | 0.7 |
| BMI (kg/m<sup>2</sup>), mean (SD) | 32.0 (7.6) | 41.3 (8.0) | - |
| Obesity class |  |  |  |
| Class I | 1 321 118 (63.2) | 2 407 (12.6) | 0.2 |
| Class II | 596 551 (28.5) | 7 239 (37.9) | 1.2 |
| Class III | 172 798 (8.3) | 9 451 (49.5) | 5.5 |
| Race and ethnicity, n (%) |  |  |  |
| Hispanic/Latino | 512 698 (24.5) | 3942 (20.6) | 0.8 |
| Non-Hispanic Asian | 51 266 (2.5) | 374 (2.0) | 0.7 |
| Non-Hispanic Black | 432 833 (20.7) | 4354 (22.8) | 1.0 |
| Non-Hispanic White | 982 474 (47.0) | 9456 (49.5) | 1.0 |
| Others | 66 257 (3.2) | 584 (3.1) | 0.9 |
| Missing | 44 939 (2.1) | 387 (2.0) | 0.9 |
| Overall SVI quartile, n (%) |  |  |  |
| Q1 (Least vulnerable) | 275 152 (13.2) | 3 242 (17.0) | 1.2 |
| Q2 | 379 578 (18.2) | 3 710 (19.4) | 1.0 |
| Q3 | 506 155 (24.2) | 4 649 (24.3) | 0.9 |
| Q4 (Most vulnerable) | 904 742 (43.3) | 7 220 (37.8) | 0.8 |
| Missing | 24 840 (1.2) | 276 (1.4) | 1.1 |
| Urbanicity, n (%) |  |  |  |
| Metropolitan | 1 740 935 (83.3) | 16 006 (83.8) | 0.9 |
| Micropolitan | 184 975 (8.8) | 1 582 (8.3) | 0.9 |
| Small towns | 88 584 (4.2) | 805 (4.2) | 0.9 |
| Rural area | 54 977 (2.6) | 453 (2.4) | 0.8 |
| Missing | 20 996 (1.0) | 251 (1.3) | 1.2 |
| Preferred language, n (%) |  |  |  |
| English | 1 840 696 (88.1) | 17 408 (91.2) | 0.9 |
| Spanish | 209 381 (10.0) | 1 424 (7.5) | 0.7 |
| Others | 40 390 (1.9) | 265 (1.4) | 0.7 |
| Insurance type, n (%) |  |  |  |
| Commercial | 737 390 (35.3) | 9 273 (48.6) | 1.3 |
| Medicaid | 903 996 (43.2) | 7 240 (37.9) | 0.8 |
| Medicare | 3 981 (0.2) | 37 (0.2) | 0.9 |
| Self-pay | 44 277 (2.1) | 45 (0.2) | 0.1 |
| Unmapped/Missing | 400 823 (19.2) | 2 502 (13.1) | 0.6 |

Summary statistics of the eligible adolescent is based on patients who meet the eligible criteria within the same period with the patients prescribed GLP1RA (January 2021–July 2025). Proportion was calculated as the number of patients prescribed GLP1RA products over the eligible population within each characteristic group (by row). Obesity class was defined as Class I if it was $\geq 100\%$ to $<120\%$ of the 95% percentile, Class II if it was between BMI $\geq 120\%$ to $<140\%$ of the 95th percentile or BMI $\geq 35$ to $<40$ , and Class III if it was 140% or more or BMI $\geq 40$ . GLP-1RA: glucagon like peptide-1 receptor agonist; Others in the Race and ethnicity included American Indian or Alaska Native, Native Hawaiian, and Pacific Islander and recorded as 'Others' in the EHR. Urbanicity was classified based on Rural and Urban Commuting Area code. 'Unmapped/Missing' in the Insurance type included a payer or payer-like entity that does not yet have a released reference payer value. SD: standard deviation; n: number; BMI: body mass index (kg/m<sup>2</sup>); SVI: social vulnerability index.

**Figure.**
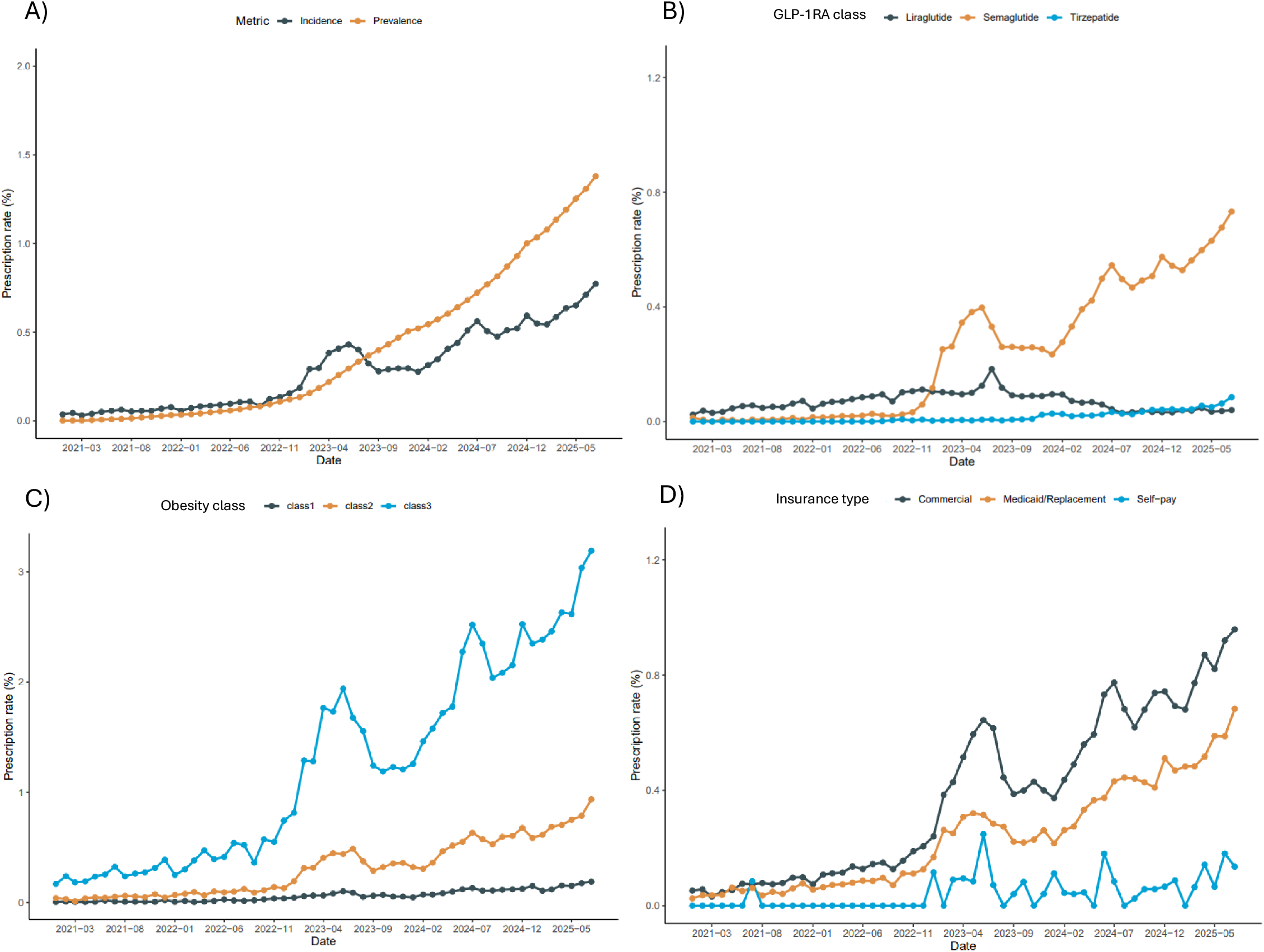
Prescription Rates of GLP-1RA in adolescents with obesity, overall and by obesity severity, GLP-1RA, and insurance type. A) Incident and prevalent prescription rate of GLP-1RA, B) Incident prescription rate by GLP- 1RA class, C) Incident prescription rate by obesity class, and D) Incident prescription rate by insurance type. All prescription rates are unadjusted rate. Obesity class was defined as Class I if it was ≥100% to <120% of the 95% percentile, Class II if it was between BMI ≥120% to <140% of the 95th percentile or BMI ≥ 35 to <40, and Class III if it was 140% or more or BMI ≥ 40. In panel d, patients with Medicare coverage were excluded due to the small sample size. GLP1-RA: glucagon like peptide-1 receptor agonist.

