## Supplemental Methods for "Prescribing of GLP-1 Receptor Agonists for Adolescents with Obesity and Associated Disparities"

**Supplementary Methods**

**Data Details**

General description

The Epic Cosmos dataset is a collaborative nationwide repository encompassing 300 million patients from over 280 US healthcare systems, linked with geospatial socioeconomic data (numbers are at the time of analysis; The dataset is continuously growing). The Cosmos dataset is a pooled dataset of electronic health records from institutions that agreed to shared governance and data use agreement documents.

While it does not comprehensively capture all patient records—since data from non-Epic healthcare systems and providers outside these participating institutions are not included—it represents a large, nationwide sample spanning healthcare systems across all 50 states (please see Figure below).

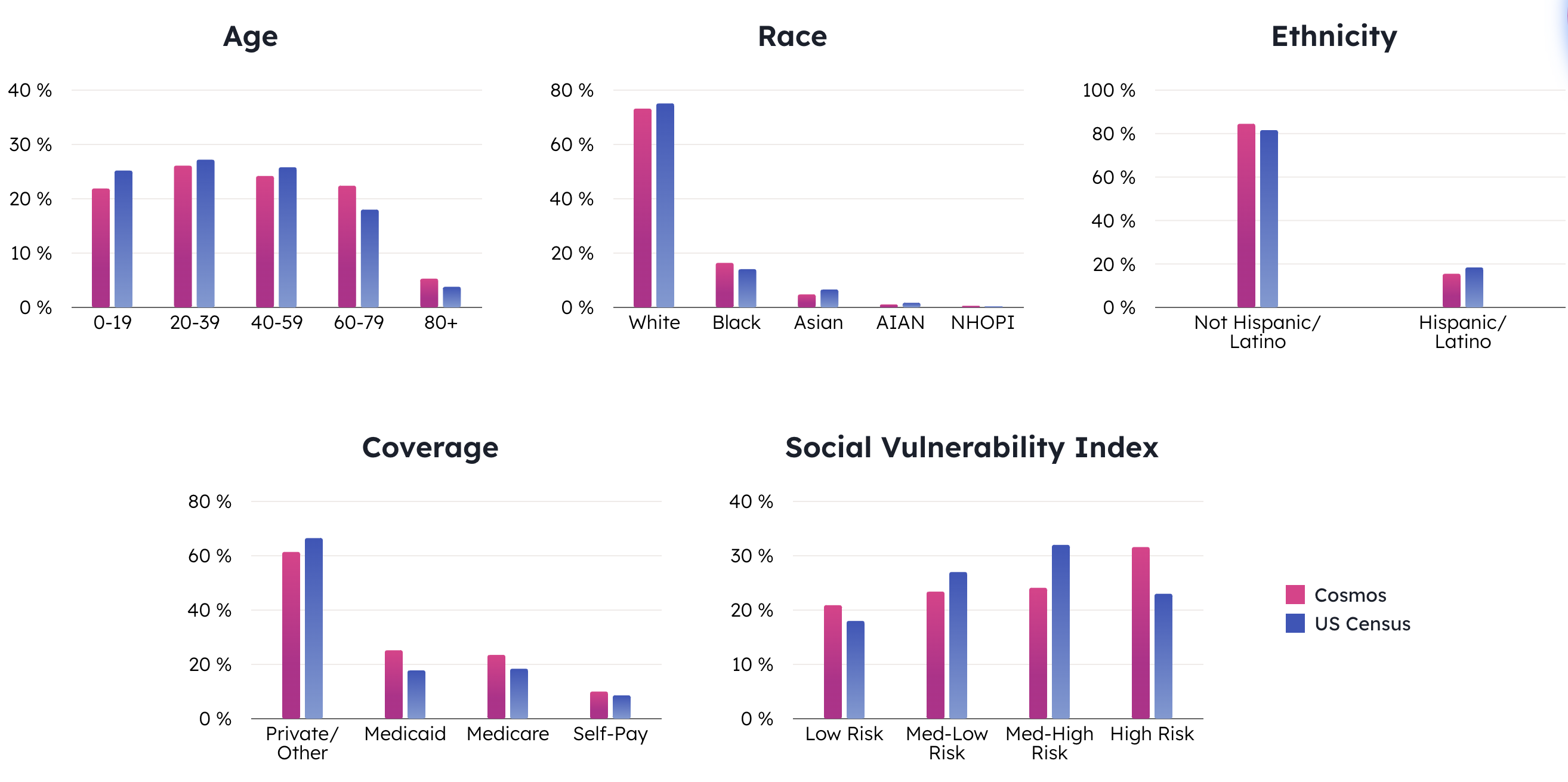

(Epic Cosmos webpage, https://cosmos.epic.com/about/)

Description for data elements

For patient information, according to the HIPAA rules, all protected health information was not contained in the Cosmos dataset. Records of the same patient across multiple institutions were de-duplicated using the same identifier.

All clinical dates in the Cosmos dataset were shifted with a randomly assigned number (0-30 days) for each patient during de-identification process by Epic.

Information on all types of encounters in the Epic system are included. This includes not only face-to-face visits such as hospital encounters and office visits, but also telemedicine, home care visits, pharmacy visits, lab visit, consultations, and so on that occur within the institution.

Medication information is based on prescription orders by healthcare systems, including elements such as brand name, generic name, type, route, quantity, units, and number of refills. However, it does not include information on pharmacy fill.

The Social Vulnerability Index (SVI) was derived from the 2018 and 2020 SVI by the Centers for Disease Control. Rural-Urban Commuting Area (RUCA) was derived from the 2010 RUCA code by the U.S. Department of Agriculture. The SVI and RUCA information were linked via the ZIP code of patients’ residence, and data linkages were made by Epic.

Clinical records in text form are not provided to researchers.

**Definitions**

Study Population

Adolescents aged 12 to 17 years with obesity, defined as body mass index (BMI) at or above the 95th percentile for sex- and age-based growth charts by the U.S. Centers for Disease Control and Prevention. Patients with a diagnosis of type 1 (ICD-10-CM E10.x) or type 2 (E11.x) diabetes in a year before the index date were excluded. Patients with incomplete BMI information were also excluded.

To ensure adequate longitudinal data and consistent engagement with the healthcare system, patients were required to have a minimum observation period of a year prior. Specifically, eligible patients must have had an initial in-person encounter (e.g., clinic or outpatient visit) documented in the Epic Cosmos dataset at least one year before the BMI measure date. Additionally, patients were required to have had at least one in-person encounter in the last year before the index date, confirming ongoing interaction with the healthcare system.

GLP-1RA

Semaglutide, liraglutide and tirzepatide were identified based on the prescription order records in the dataset. Prescription records from other countries (Canada, Lebanon, and Saudi Arabia) than the U.S. were not considered for this study. Given the high off-label use of products for diabetes to manage obesity, we included not only obesity-specific brands (Wegovy®, Saxenda®, and Zepbound®) but also diabetes-specific brands (Ozempic®, Victoza®, and Mounjaro®) however, excluded patients with a diagnosis code of type 2 diabetes. Oral product (Rybelsus®) and insulin-combined product (Xultophy®) was not considered for this study.

Race and Ethnicity

Race and ethnic groups were classified as non-Hispanic White, non-Hispanic Black, Hispanic, non-Hispanic Asian, and Others. If the Ethnicity column in the dataset specified Hispanic or Latinos, patients were defined as the Hispanic group; otherwise, those were classified as Non-Hispanic White, Non-Hispanic Black, and non-Hispanic Asian based on the First Race column. The Others group included American Indian or Alaska Native, Native Hawaiian, and Pacific Islander and recorded as ‘Others’. If there was no race information, it was considered missing.

Obesity Class

Obesity class was determined based on adolescents’ BMI. Class I obesity defined as if BMI was ≥100% to <120% of the 95% percentile for sex- and age-based growth charts by the U.S. Centers for Disease Control and Prevention, Class II was if it was between BMI ≥120% to <140% of the 95th percentile or BMI ≥ 35 to <40, and Class III was if it was 140% or more or BMI ≥40.

Insurance Information

Insurance type was determined based on Reference Financial Class information from Cosmos. The financial class represents the category of payer associated with the encounter, such as commercial insurance, Medicare, or Medicaid. Encounter table contains information about financial class and whether the encounter was marked as self-pay.

‘Unmapped/Missing’ in the table for Insurance type indicates a payer or payer-like entity that does not yet have a released reference payer value. This is distinctive from other values in that this group is intended to be temporary. For example, a new commercial payer may exist without a mapping for some time. Once it that payer was given a value, it may then be updated on Cosmos as a Commercial payer.

Prescription rate

Prescription rate was calculated in two different manners, incident prescription rate or prevalent prescription rate. The definitions of two metrics were based on previous literature^1^. Duration of prescription didn’t affect the calculation of the incident or prevalent prescription rates.

1) Incident prescription rate

The numerator of the incident prescription rate was calculated as the number of eligible patients who received their first ever prescription of GLP-1RAs in a given month. The denominator was calculated as the number of eligible patients who had any medical records but had never been prescribed GLP-1RAs before in a given month.

$$Incident prescription rate= \frac{Number of eligible patients received the first prescription}{Number of eligible patients who never be prescribed before}$$

For example, the incident prescription rate in June 2021 was calculated as (the number of eligible patients who received their first ever semaglutide or tirzepatide prescription in June 2021) over (the number of eligible patients who had never prescribed GLP-1RAs before June 2021).

2) Prevalent prescription rate

Due to the nature of observational data, it is difficult to calculate the true denominator of prevalence. Therefore, we defined the prevalent prescription rate as 1-year running prevalence. The numerator of the prevalent prescription rate was calculated for each month as the number of eligible patients who have received GLP-1RAs prescriptions in the previous 12 months. The denominator was calculated for each month as the number of eligible patients who had any medical records in the database over the past year.

$$Prevalent prescription rate= \frac{Number of eligible patients received a prescription in the previous year}{Number of eligible patients having any records in the database over the past year}$$

For example, the prevalent prescription rate in June 2021 was calculated as (the number of eligible patients who received any GLP-1RAs prescription between July 2020 and June 2021) over (the number of eligible patients who had any medical record between July 2020 and June 2021).
